# Prediction on Covid-19 epidemic for different countries: Focusing on South Asia under various precautionary measures

**DOI:** 10.1101/2020.04.08.20055095

**Authors:** Abhijit Paul, Samrat Chatterjee, Nandadulal Bairagi

**Author notes:** corresponding authors (NB), (SC).

## Abstract

The coronavirus disease 2019 (COVID-19), which emerged from Wuhan, China, is now a pandemic, affecting across the globe. Government of different countries have developed and adopted various policies to contain this epidemic and the most common were the social distancing and lockdown. We proposed a SEIR epidemic model that accommodates the effects of lockdown and individual based precautionary measures and used it to estimate model parameters from the epidemic data up to 2nd April, 2020, freely available in GitHub repository for COVID-19, for nine developed and developing countries. We used the estimated parameters to predict the disease burden in these countries with special emphasis on India, Bangladesh and Pakistan. Our analysis revealed that the lockdown and recommended individual hygiene can slow down the outbreak but unable to eradicate the disease from the society. With the current human-to-human transmission rate and existing level of personal precautionary, the number of infected individuals in India will be increasing at least for the next 3 months and the peak will come in 5 months. We can, however, reduce the epidemic size and prolong the time to arrive epidemic peak by seriously following the measures suggested by the authorities. We need to wait for another one month to obtain more data and epidemiological parameters for giving a better prediction about the pandemic. It is to be mentioned that research community is working for drugs and/ or vaccines against COVID19 and the presence of such pharmaceutical interventions will significantly alter the results.

## 1 Introduction

A local outbreak of pneumonia was detected in Wuhan city of Hubei Province, China, in December 2019, which was later known to be caused by a novel coronavirus, severe acute respiratory syndrome coronavirus 2 (SARS-CoV-2)^1^ and China became the epicenter^2^. This communicable disease has symptoms like fever, dry cough, breathlessness and fatigue^3^. With human migration, the disease has now spread over 206 countries or territories of the world, making Europe and USA as new epicenters^4,5^. SARS-CoV-2, which causes coronavirus disease 2019 (COVID-19), has been given pandemic status by the World Health Organization (WHO) on 11th March, 2020^6^. With 9,00,306 confirmed cases and 45,693 confirmed death^7^ as on 2nd April, 2020, COVID-19 has already exceeded the previous records of two coronavirus epidemics (severe acute respiratory syndrome coronavirus, SARS-CoV, and Middle East respiratory syndrome coronavirus, MERS-CoV), posing the biggest threat to the global public health and economy after the second world war^8^.

Due to unavailability of pharmaceutical interventions, government of different countries are adopting various policies to contain this epidemic and the most common is lockdown. It started with the local government of Wuhan by suspending all public traffics within the city on 23rd January 2020 and soon followed by other cities in Hubei province^9^. In the absence of drug and vaccine for this disease, maintaining social distancing is the only way to reduce person-to-person transmission of COVID-19^10^, and thus other countries also implemented lockdown, quarantines and curfews. In response to the growing COVID-19 pandemic, the Italian Government imposed a national quarantine on 9th March 2020^11^, restricting the movement of people except for necessity and emergency health care. It was followed by Spain (14th March 2020^12^), Argentina (19th March 2020^13^), United States (different states from 19th March, 2020^14^), United Kingdom (23rd March, 2020^16^), South Africa (26th March^15^) and many other countries.

Like rest of the world, South Asian countries also started taking measures to fight against the rapid spread of COVID-19. South Asian leaders met over video conference on 15th March 2020 to discuss on this issue and proposed to create a common fund classified as COVID-19 Emergency Fund for the SAARC (South Asian Association for Regional Cooperation) countries^17^. In India, the first case was reported in Kerala on 30th January 2020 when a student returned back from Wuhan^18^. Indian Government implemented a complete lockdown across the country on and from 25th March for 21 days, following one day ‘Janata Curfew’ on 22nd March, to prevent COVID-19 epidemic in India^19^. Pakistan confirmed its first two cases of the coronavirus on 26th February, 2020^20^, but yet to go for complete lockdown^23^. Institute of Epidemiology, Disease Control and Research (IEDCR) confirms the first three known cases of COVID-19 in Bangladesh on 7 March 2020^21^. The country announced one week shutdown on March 26 and later on extended it till 11th April^22^.

The aim of the present study is to predict the course of COVID-19 epidemic in different countries based on the current epidemic trends observed in those countries. We also want to analyze how the effect of various government measures taken at the national level like complete lockdown; measures at community level like social distancing; measures at individual level like using of mask and hand-wash, have on the incidence of new cases in different countries affected by COVID -19. Finally, the study focuses on the possibility of disease control and eradication in the South Asian countries.

## 2 Method and Results

### 2.1 Country-wise COVID-19 Outbreak Data

The data used for the current study is freely available in GitHub repository^24^, along with the feature layers of the dashboard, which are now included in the Esri Living Atla^1^. This is the data repository maintained by the Johns Hopkins University^24,25^. In the current study, we have used the country-wise temporal data of confirmed, recovered and death cases till 2nd April, 2020. The data is represented in a time series graph given in the supplementary Fig. S1. For the current study, we have not considered the data from China because a lots of study has already been done^26,27^ and, moreover, the epidemic curve of China has already passed its peak^24^, while the curve for other countries are still growing up and therefore prediction for such countries might be helpful for the policy makers and health care service providers in curbing the spread of this dreaded disease.

### 2.2 The general mathematical model

We proposed here a general mathematical model that can represent the overall dynamics of COVID-19 epidemic. The entire human population of a country was classified into four compartments, viz. susceptible population (who are supposed to be infected by the disease), exposed population (who have contacted the disease but not yet infectious), infective population (who are capable of spreading the disease), recovered population (who have recovered from the disease and no more infectious) to construct a SEIR (susceptible (*S*) → exposed (*E*) → infectious (*I*) → recovered (*R*)) model. The model was further extended to accommodate a death class (*D*) (who have died due to this disease) for assessing the reported death^28^ due to coronavirus. The rate of change in each compartment at time *t* is represented by the following set of differential equations,

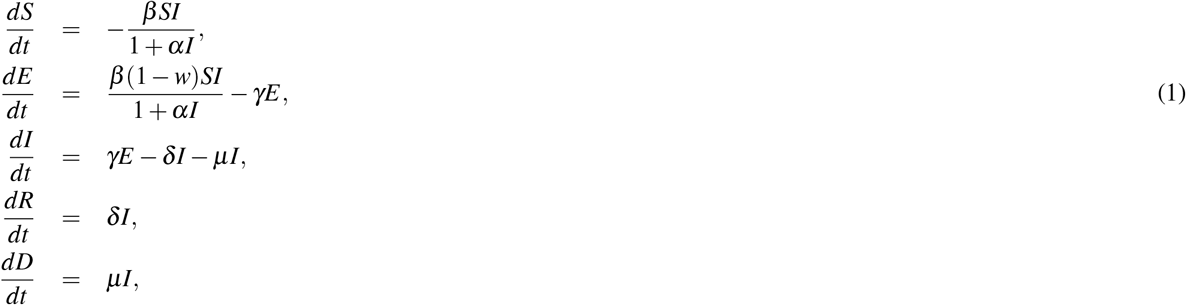

where the transmission efficiency is represented by *β* = *ne*, where *n* is per capita daily number of contacts per unit of time and *e* is the probability of transmission per contact between a susceptible and an infective^28^. The effect of the parameter *β* is directly related with the measures like lockdown, restriction of movement, shaking hand etc. which actually reduce the number of contacts. While the parameter *w*(0 < *w* < 1) encapsulates the efficacy of various individual precautionary measures, like cough etiquettes, frequent washing of hand with soap and sanitizer etc. Usually, incidence rate (the rate at which susceptible become infective) in epidemiological models is considered as bilinear (proportional to the product of *S* and *I*) type, also called mass-action type. This assumption is valid when there is a small number of infective, however, this law fails as the infective numbers increases^29^, because the per capita contacts decreases with increasing *I*. It is therefore assumed that the disease incidence is of nonlinear and of mass-action saturated type, which behaves like bilinear type incidence when *I* is small and saturates as *I* increases^30,31^. It is also shown that this nonlinear incidence better fit the epidemiological data compared to bilinear incidence^32^. The nonnegative parameter, *α* measures the extent of physiological effect. Exposed individuals progress to infectious class at a rate *γ*, thus the average time spent in the exposed class is 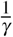 unit of time. Infectious individuals either recover at a rate *δ* or die at a rate *µ*, implying the mean infectious period is 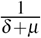 unit of time. The total population, *N*, is assumed to be constant for a designated area so that normal birth and death balance each other in the study period.

### 2.3 Country-wise parameter estimation

For this analysis, we selected countries from different geographical locations like Europe (Italy, Spain, Germany, France and United Kingdom), North America (USA) and South Asia (India, Pakistan and Bangladesh). We use ODE45 and curve fitting toolbox of MATLAB to estimate the parameter values that best fit the real time data^24,25^. The estimated curves for country-wise cumulative confirmed cases are presented in Fig. 1 (similar figures for recovered and death cases are presented in supplementary Fig. S2 and Fig. S3), which evidently represents the real data of COVID-19 infection. The model parameters corresponding to the best fit curve for different countries are presented in Table 1, which will be further used to make different predictions.

**Table 1.** Country-wise estimated parameter values for the model system (1). The population for each country *N* is taken from literature.

| Parameters | $N$<br>(individual) | $\beta$<br>(per day per individual) | $w$<br>($0 < w < 1$) | $\alpha$<br>(per individual) | $\gamma$<br>(per day) | $\delta$<br>(per day) | $\mu$<br>(per day) |
| --- | --- | --- | --- | --- | --- | --- | --- |
| US | 32,92,27,746<sup>33</sup> | 1.1 | 0.8 | 1.2 | 0.5151 | 0.0062 | 0.004 |
| Italy | 6,03,17,546<sup>34</sup> | 1.21 | 0.787 | 1.2 | 0.81 | 0.034 | 0.026 |
| Spain | 4,67,33,038<sup>35</sup> | 1.26 | 0.767 | 1.2 | 0.851 | 0.0591 | 0.0225 |
| Germany | 8,30,42,200<sup>36</sup> | 1.13 | 0.79 | 1.2 | 0.624 | 0.052 | 0.0025 |
| France | 6,70,22,000<sup>37</sup> | 1.2 | 0.82 | 1.2 | 0.58 | 0.038 | 0.016 |
| United Kingdom | 6,78,86,004<sup>38</sup> | 0.9333 | 0.82 | 1.2 | 0.669 | 0.00081 | 0.012 |
| India | 135,26,42,280<sup>39,40</sup> | 0.1711 | 0.1 | 1.2 | 0.45 | 0.0091 | 0.0034 |
| Pakistan | 21,22,28,286<sup>41,42</sup> | 0.281 | 0.1 | 1.2 | 0.52 | 0.01 | 0.00265 |
| Bangladesh | 16,13,76,708<sup>41,42</sup> | 0.1955 | 0.1 | 1.2 | 0.455 | 0.079 | 0.0187 |

**Figure 1.**
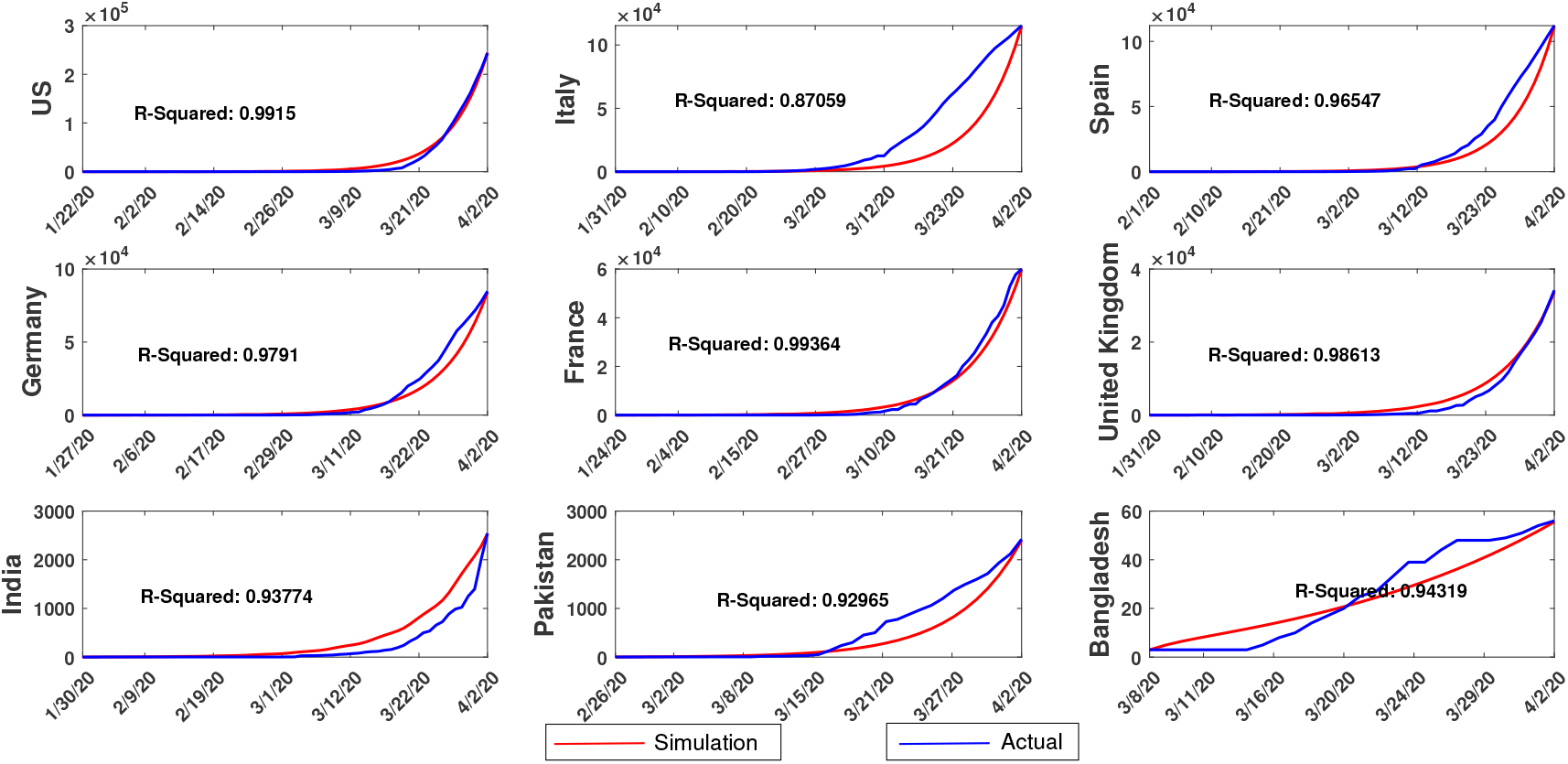
Cumulative confirmed cases: Comparison of country-specific simulation results of our proposed model (1) with the published data for confirmed cases. The R-squared value for each case is mentioned in the inset. The data contains information on daily basis from 22nd January to 2nd April, 2020. To keep the figure clean we have only marked some of the dates.

A comparison among the model parameters of different countries are given in Fig. 2. It shows that the efficacy of various individual level measures, like washing of hand and using of mask, is more in USA and Europe than in South Asian countries. This is expected because of the difference in the socio-economic conditions of these countries. But, it was interesting to note that the daily contact (*n*) is low in developing nation in comparison to developed countries with India at the lowest and Spain at the highest position. One probable reason for such result is that India announced lockdown much earlier than USA and other countries after the detection of first positive case. This restricted the person-to-person transmission significantly in India, breaking the chain. The trend, however, may change if violation of lockdown occurs and/or detection processes is not rapidly done. As expected, the conversion rate of the exposed class to infected class is highest in Italy and Spain. It is lowest in India and Bangladesh because the epidemic is at initial stage in these countries. The recovery rate is also observed to be different in various countries and surprisingly lowest in USA and UK.

**Figure 2.**
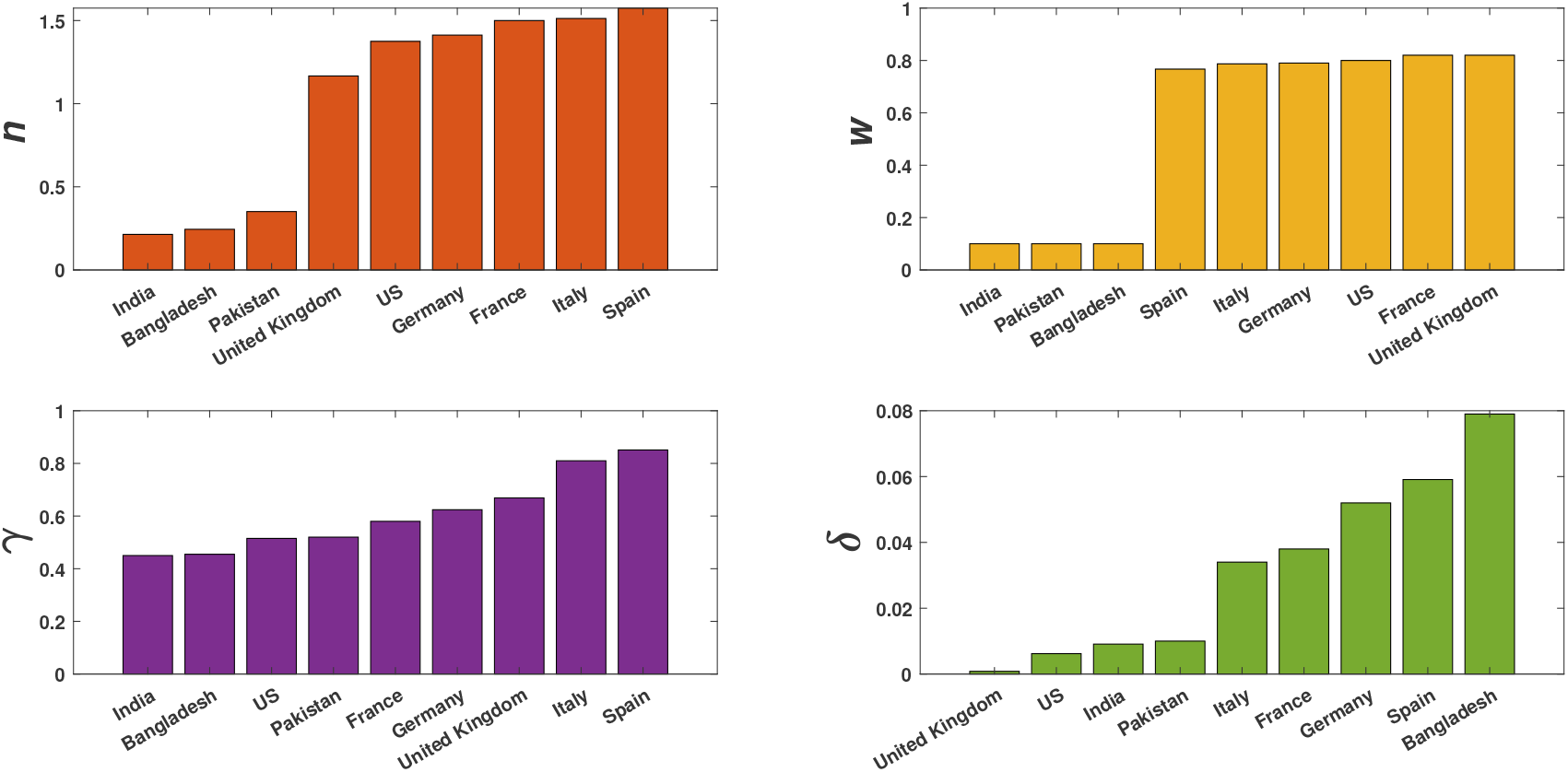
Country-wise comparison of important parameters estimated from our model: The value of *n* was obtained from the relation *β* = *ne* with *e* = 0.8. The value of *e* was assumed to be high as the probability of person-to-person transmission for this disease is significantly high. The other parameters are as defined in the Table 1.

### 2.4 Effect of different parameters on epidemic progression of different countries

After estimating the model parameters for different countries based on the real data, we studied the significance of different parameters on the course of epidemic. We considered two parameters *β* and *w*, which are most affected by lockdown and individual level precautionary measures, and studied their influence on the epidemic curve. We varied the said parameters and calculated the fold change in infected, recovered and death cases of each country after one month, considering the data of 2nd April as their respective base values. Here we have presented the results of USA and Italy (Fig. 3), and India (Fig. 4). Similar results for rest of the countries are given in the supplementary (Figs. S4-S6). All the countries showed a declined trend in infected numbers with the decreasing rate of human-to-human transmission, however, the rate differed from country to country. The individual precautionary measures, *w*, is high for the European countries and USA and so for these countries there is little scope for any further increase in *w*. However, the estimated value of *w* for South Asian countries is too low and there is a huge scope for increasing the value of *w* with good hygiene practice.

**Figure 3.**
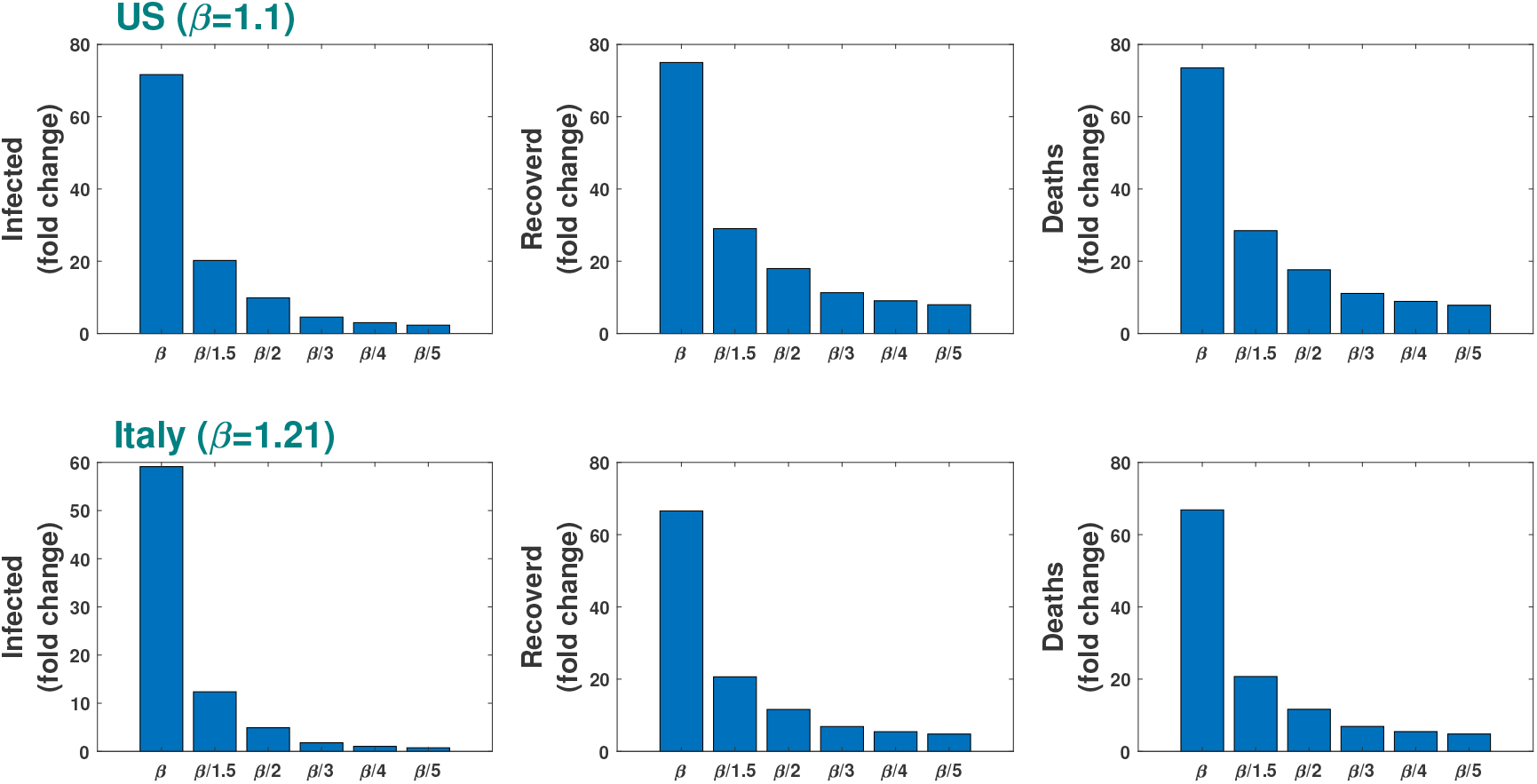
Effect of parameter variation: Estimated fold change in the number of infected, recovered and death cases for COVID-19 epidemic in USA and Italy after one month for different values of *β* (upper row) and *w* (lower row). The fold change on 3rd May was calculated on the basis of respective numbers of 2nd April.

**Figure 4.**
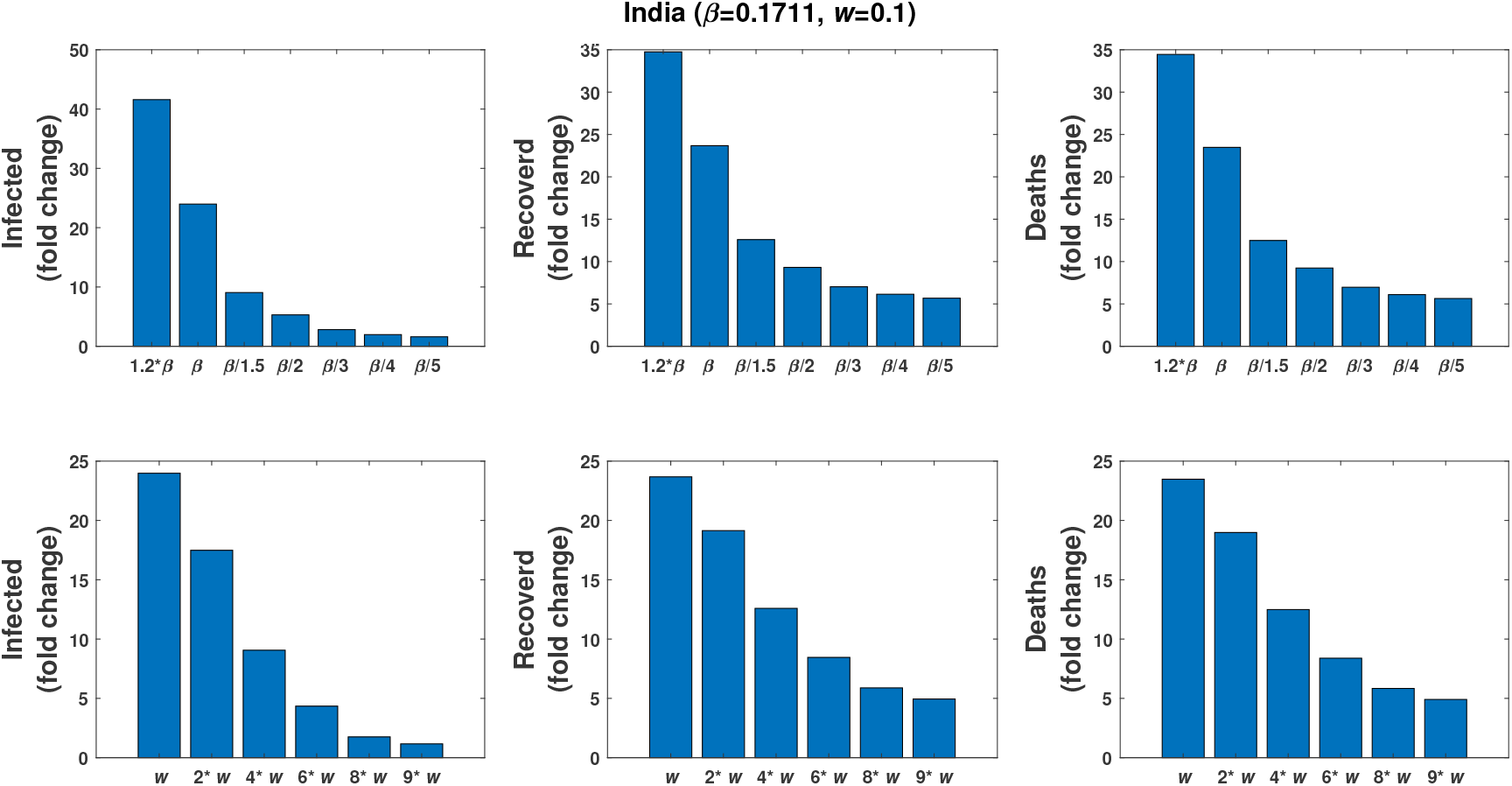
Effect of parameter variation: Estimated fold change in the number of infected, recovered and death cases for COVID-19 epidemic in India after one month for different values of *β* (upper row) and *w* (lower row). The fold change on 3rd May was calculated on the basis of respective numbers of 2nd April 2020.

In the case of India also a decreasing trend is observed in the numbers of infective, recovered and death cases due to variation in *β* and *w*. If the estimated value of *β* is maintained (see Table 1), the fold change after one month in the respective classes will be 30, 22 and 24 times of the base value and this will become 9, 8, 14 times if the value of *β* is reduced to halved of its current estimated value due to lockdown effect. If for some reason *β* becomes higher (1.2 times of the current value), e.g., in the case of “superspreading” (when a single infected individual infects a large number of susceptible^28^), this values becomes 42, 35, 34 times. Similar results for variation in *w* are plotted in the lower row of Fig. 4.

The prevalence in epidemiology, defined as the proportion of infected individuals in the total population^43^, is used to indicate the severity of the disease. It also quantifies the chance of transmissibility, i.e., with higher prevalence value there is more chances of disease transmission. We plotted disease prevalence of different countries with the estimated parameter values and predicted the disease load for the next 6 months in Fig. 5. It shows that disease prevalence declines significantly and hence the chances of person-to-person transmission also reduces in all European countries except UK. Disease prevalence status is almost similar in the case of UK and USA.

**Figure 5.**
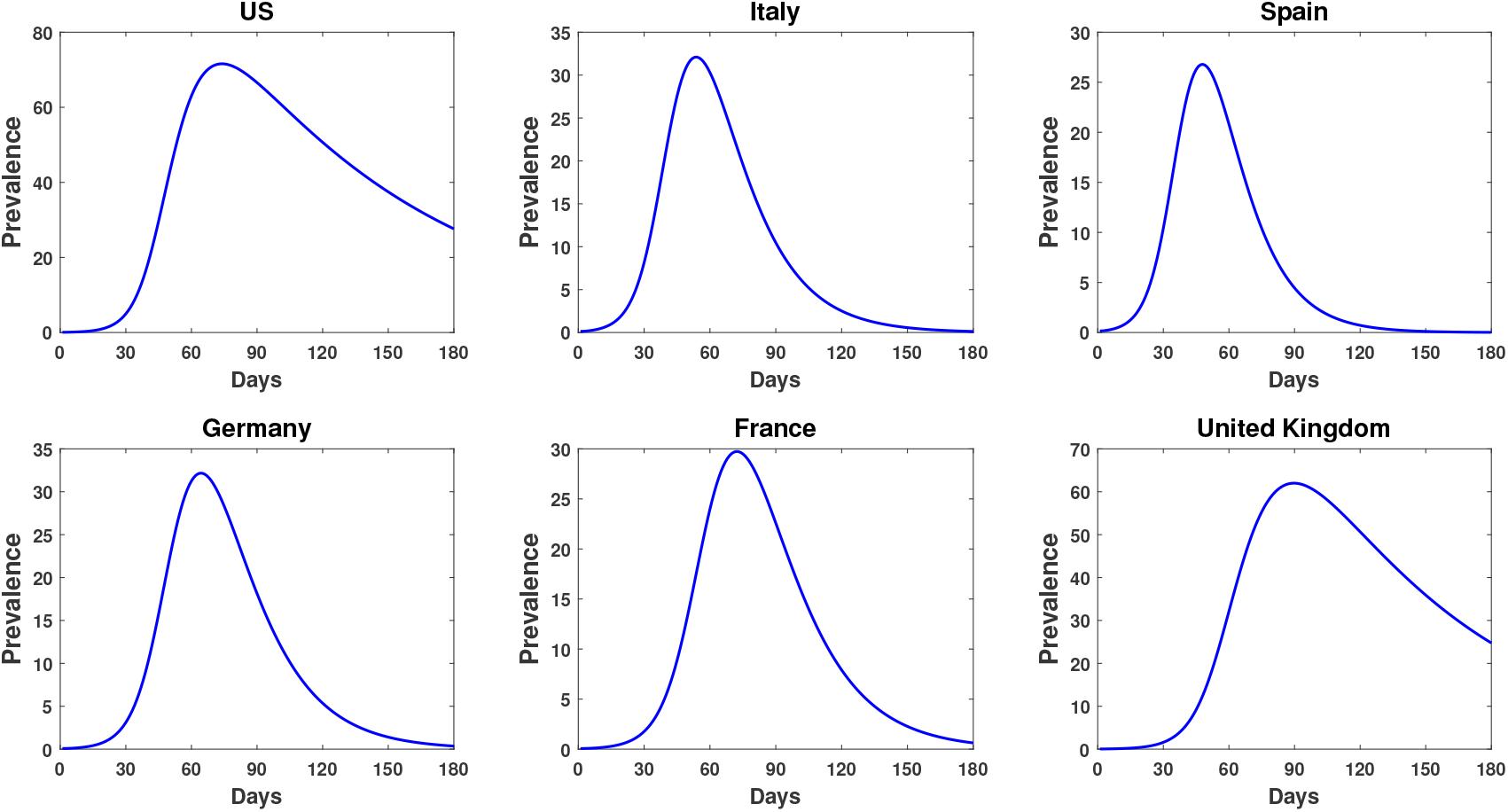
Country-wise prevalence percentage. 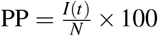 for different countries was plotted using the estimated parameter values as in Table 1, where *I*(*t*) is the infected population at time *t* an *N* is the total population of that country.

In an unmitigated scenario, the prevalence results (Fig. 6) on South Asian countries predicted that the number of infected individuals in Pakistan will reach its peak in 85 days with 70% of the total population gets infection. In India, the peak will reach in 150 days with 60% infected population. The situation is quite different in Bangladesh, where increasing trend sustains for much longer time. The peak in Bangladesh will come in around 230 days at its current transmission rate and it will affect around 8% of its population. We want to mention here that the prediction on Bangladesh may drastically change because the current available data is very low.

**Figure 6.**
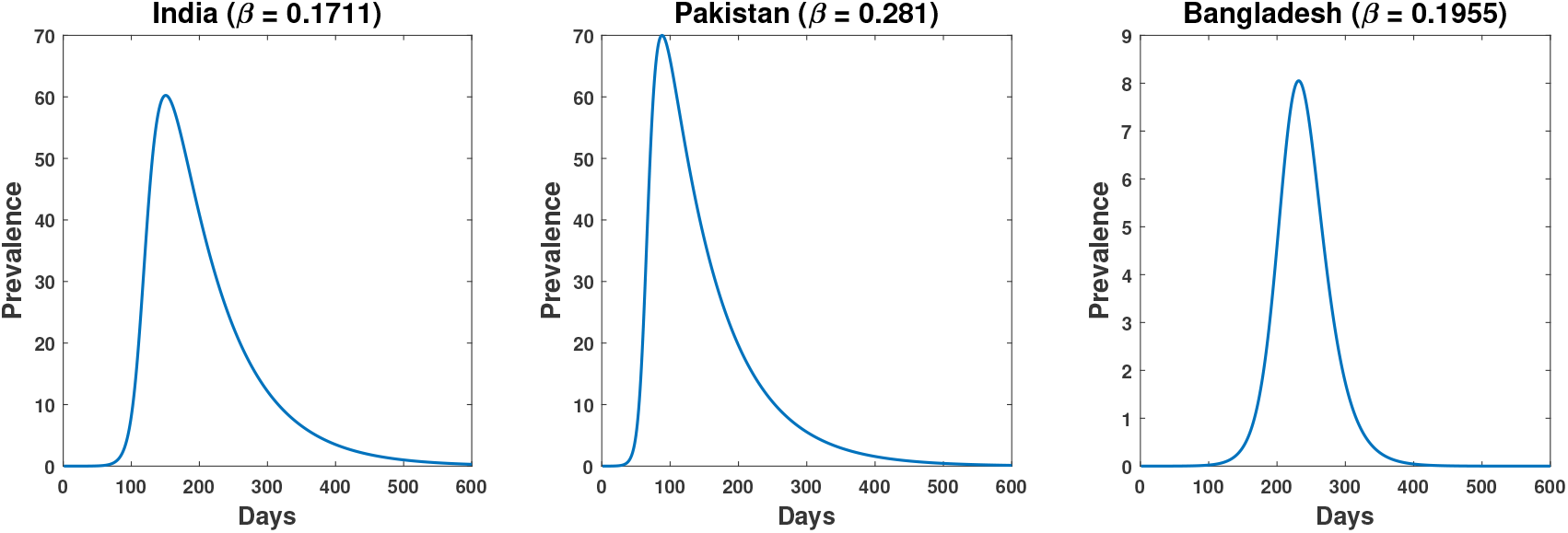
Prevalence percentage of South Asian countries. This figure shows that abut 60% of Indian population will be infected with the coronavirus after 150 days if disease progression occurs with the estimated force of infection. Pakistan will face the highest disease burden (70%) after 85 days. Disease prevalence will be lowest in Bangladesh, which is only 8%, but this value may vary drastically as the present data available for Bangladesh is very low.

### 2.5 Prediction on control measure for South Asian countries

To predict on the outcomes of control measures in the South Asian countries, we studied the prevalence percentage for India and Pakistan by decreasing the infection force, *β*, via reduced contact rate (*n*) and increasing the individual hygienic measure, *w*. If the outbreak continues with the respective estimated *β* value, India’s disease prevalence percentage after 90 days will be 3% (approximately) of the total population and the same for Pakistan is 70% (see Table 2). Moreover, if *β* is decreased by 2 fold, i.e. if the human-to-human transmission rate is reduced by 50% of the estimated rate due to lockdown, then disease prevalence in India would decrease to 0.026%, while for Pakistan it will be still above 4%. To decrease the prevalence percentage in Pakistan below 0.3%, human-to-human transmission rate has to be decreased by 66% of the present rate (Table 2). Based on the available report, Bangladesh is showing the best control over the disease spread in comparison to India and Pakistan. Similar results for *w* are also presented there.

**Table 2.**
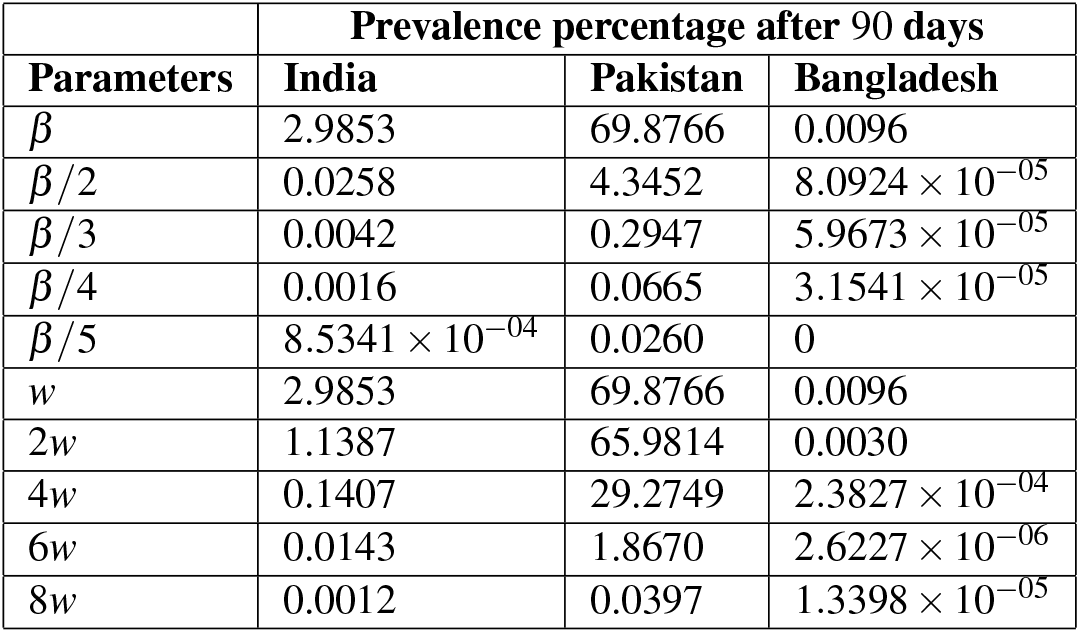
Changes in the prevalence of disease (after 90 days) with the variation of *β* and *w*.

Another interesting result which we observed as an effect of lockdown is in the occurrence of epidemic peak. Lockdown not only reduces the size of epidemic but also causes delay in the occurrence of peak (Fig. 7). For example, if lockdown effect reduces the existing contact rate by 50%, the peak value will reduce to 43% and will come around after 280 days. If it reduces the contact rate to one-third of the existing value, then the prevalence will be 32% and will appear after 423 days (see blue colour curve in each figure of Fig. 7). It will be further delayed with reduced disease burden if the efficacy level of individual precautionary measures is increased. For example, if the lockdown effect reduces the contact rate to 50% and individual hygiene efficacy (*w*) is increased by 4 fold then the epidemic peak will be reduced to around 32% and will appear after 423 days. Thus, these two parameters not only decrease the infected population level but also make significant delay in the appearance of peak value, allowing extra time to the authority in improving the existing healthcare systems and giving a chance to have some pharmaceutical interventions by this time.

**Figure 7.**
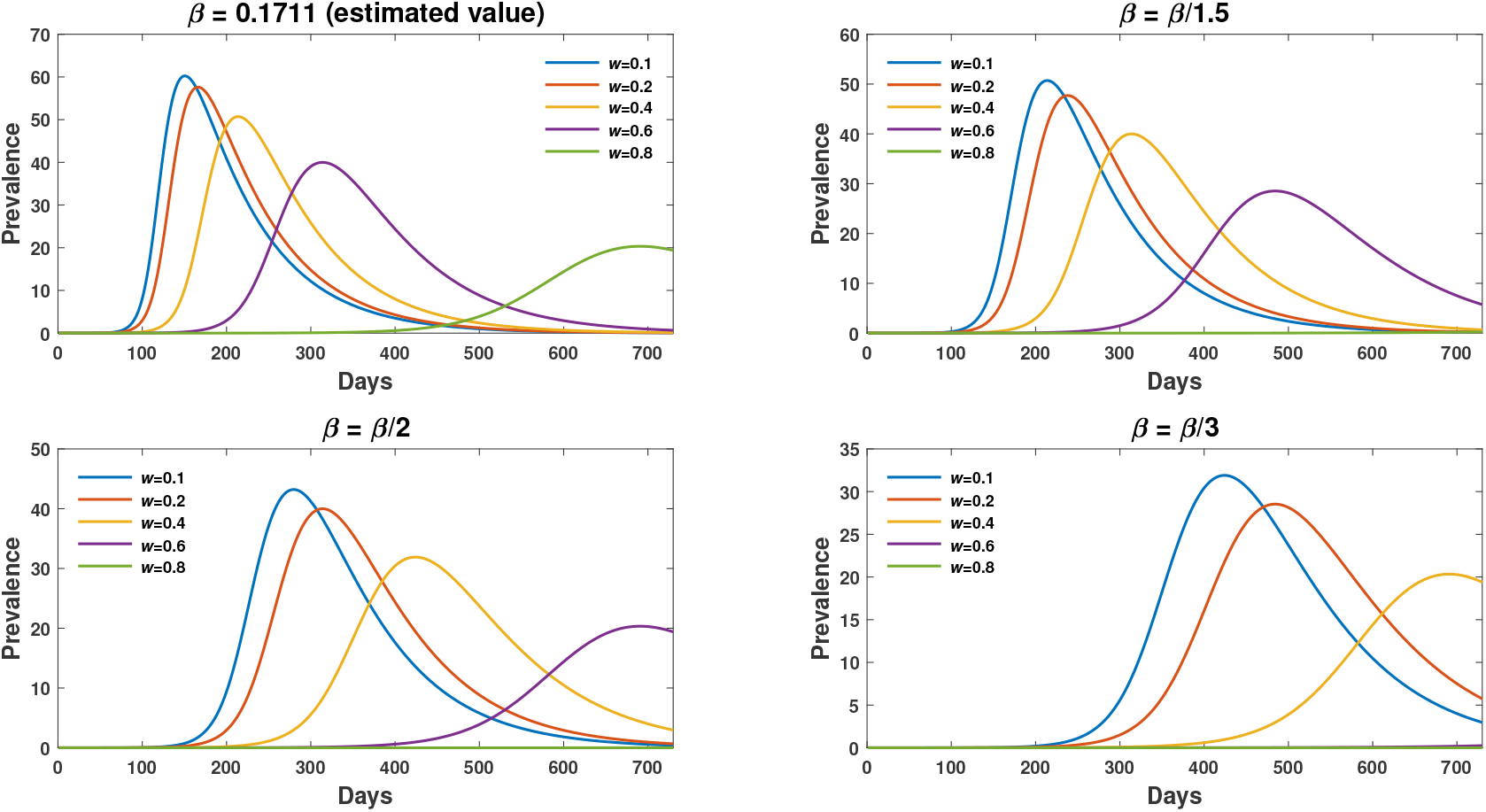
Combined effect of *β* and *w* in the prevalence percentage. Disease load in India due to the variation in the lockdown effect (*β*) and efficacy of individual precautionary measures (*w*).

We have observed that by decreasing *β* or increasing *w*, the number of infective can be reduced to some extent. We, therefore, searched for a combination of these parameters that would reduce the number of infected individual from the present number after one month. For such study, we computed a 2D parameter space (Fig. 8) to see if there exists any such combination. We obtained a region (green colour) from where if we take the parameter values then it will reduce the infected population below the current value and thus hypothesized as a threshold control measure. The red region shows the parameter combination for which the infected population will always increase from the present value. The current estimated combination of *β* and *w* is marked by a green dot.

**Figure 8.**
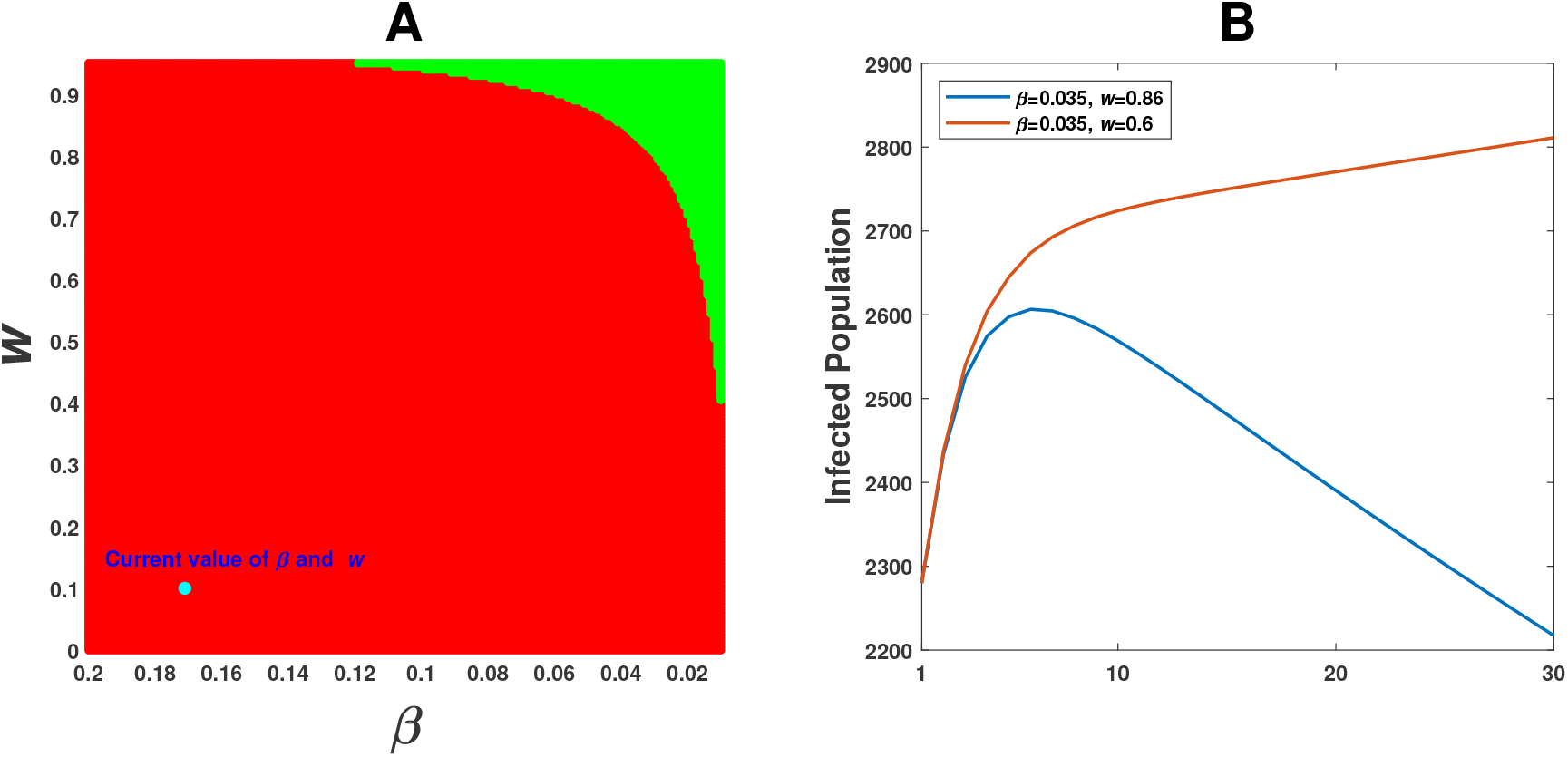
Coupling effect of parameters: For India, we plotted a 2D parameter space with *β* and *w*. The green region in the 2D space is the parameter combinations for which the infected population will significantly decrease from the present value after one month. The red region shows the parameter combination for which the infected population will increase from the present value. The cyan dot represents the current estimated value of *β* and *w*. Time series graph of infected population with two sets of *β* and *w*, taking one each from the green ad red regions, are plotted in Fig. B.

## 3 Discussion

We proposed a SEIR model and used the available data^24,25^ to estimate the parameters for the spread of COVID-19 in nine different countries. The time series data, starting from 22nd January to till 2nd April, 2020, for both the developed and developing countries were taken into consideration for the estimation of country-wise parameter values. We then varied the estimated parameter values to see their effect on the course of epidemic in each country. The effect of lockdown and personal hygiene, which affect the human-to-human transmission at the community level and household level, were considered in the model through the parameters *β* and *w*. As expected, lockdown and individual hygiene measures had shown their influences in reducing the epidemic burden in all nine countries. Looking at the country-wise prevalence percentage for the next three months, we observed that disease burden will decline in all the four considered European countries except UK. The situation of USA is almost similar to UK, where disease prevalence will be around 25%. At the current infection rate, India and Pakistan will reach their peak with disease prevalence around 60% and 70%, respectively, after 150 and 85 days. Bangladesh will attain its epidemic peak much later. The best part in Bangladesh is that the highest disease prevalence is lower than 9%, which however may be the result of small amount of data availability.

We were more interested to see the effect of lockdown and individual precautionary measures on India and Pakistan, though the latter is yet to go for complete lockdown. With the existing contact rate, the epidemic in Pakistan is going to affect around 70% of its population in the coming 3 months, where as 3% of Indian population will be affected by the disease in the same duration. However, the number of infective in India will exponentially grow in the next 2 months and the disease prevalence may be about 60% of the total population. To decrease the prevalence percentage to 0.03% in next 3 months, Pakistan has to reduce the human-to-human transmission rate to one-fifth of the current value, while for India it can be achieved by reducing it to half of the existing rate. Lockdown not only reduces the size of epidemic but also causes delay in the occurrence of peak. The length of delay increases further if the individual level precautionary measures are increased, see Fig. 7.

It is observed that the non-pharmaceutical measures, lockdown, maintaining social distance and individual hygiene, have negative correlation with infective numbers. But none of these can individually decrease the infected population from its current value. They, however, can prevent the outbreak within a period of one month when applied jointly and their measures remain above some threshold values. Our 2D parameter space analysis showed the existence of such parametric region (the green colour portion in Fig 8) even when there is no pharmaceutical intervention. Number of infected population would decrease from its current value after one month if the parameters (*β* and *w*) attain their values from the green zone. Though this region is very small, but it could be attainable if every individual strictly follows the lockdown and social distance at the community level and personal hygiene at the household level.

Finally, we want to conclude that the number of infected populations is going to increase in India for the next 3 months if human-to-human transmission and personal precautionary measure continue with the existing rates. We can, however, reduce the size of epidemic and prolong the time to arrive at the peak of epidemic by seriously following the measures suggested by the authorities. We need to wait for another one month to obtain more data and epidemiological parameters for giving a better prediction about the pandemic. It is to be mentioned that research community is working for drugs and/ or vaccines against COVID19 and the presence of such pharmaceutical interventions will significantly alter the results.

## Data Availability

The data used for the current study is freely available in GitHub repository.

https://github.com/CSSEGISandData/COVID-19

## Acknowledgements

The work is supported by SERB (Govt of India) under MATRICS Scheme, Ref no. MTR/2018/000791.

## Author contributions

All authors have equally contributed.

## Supplementary figure legends

**Figure S1. Available data on COVID-19.** The data from the period of 22nd January, 2020 to 2nd April, 2020. (a) Confirmed cases, (b) Recovered cases, (c) Death cases.

**Figure S2. Cumulative recovered cases:** Comparison of country-specific simulation results of our model (1) with the published data of recovered cases.

**Figure S3. Cumulative death cases:** Comparison of country-specific simulation results of our proposed model (1) with the published data for death cases. Bangladesh does not show a good curve fit due to lack of sufficient data.

**Figure S4. Effect of parameter variation:** Estimated fold change in the number of infected, recovered and death cases for COVID-19 epidemic in Europe after one month for different values of *β*. The fold change on 3rd May was calculated on the basis of respective numbers of 2nd April 2020.

**Figure S5. Effect of parameter variation:** Estimated fold change in the number of infected, recovered and death cases for COVID-19 epidemic in Pakistan after one month for different values of *β* (upper row) and *w* (lower row). The fold change on 3rd May was calculated on the basis of respective numbers of 2nd April 2020.

**Figure S6. Effect of parameter variation:** Estimated fold change in the number of infected, recovered and death cases for COVID-19 epidemic in Bangladesh after one month for different values of *β* (upper row) and *w* (lower row). The fold change on 3rd May was calculated on the basis of respective numbers of 2nd April 2020.

